# Characterizing traumatic brain injury and its association with homelessness in a community-based sample of precariously housed adults and youth

**DOI:** 10.1101/19004606

**Authors:** Jacob L. Stubbs, Allen E. Thornton, Kristina M. Gicas, Tiffany A. O’Connor, Emily M. Livingston, Henri Y. Lu, Amiti K. Mehta, Donna J. Lang, Alexandra T. Vertinsky, Thalia S. Field, Manraj K. Heran, Olga Leonova, Tari Buchanan, Alasdair M. Barr, G. William MacEwan, William G. Honer, William J. Panenka

**Affiliations:** Department of Psychiatry, University of British Columbia, Vancouver, Canada; British Columbia Mental Health and Substance Use Services Research Institute, Vancouver, Canada; Department of Psychology, York University, Toronto, Canada; Department of Psychology, Simon Fraser University, Burnaby, Canada; Department of Radiology, University of British Columbia, Vancouver, Canada; Division of Neurology, University of British Columbia, Vancouver, Canada; Department of Anesthesiology, Pharmacology, and Therapeutics, University of British Columbia, Vancouver, Canada; British Columbia Neuropsychiatry Program, University of British Columbia, Vancouver, Canada

**Keywords:** traumatic brain injury, homeless, precarious housing, marginally housed

## Abstract

We characterized the prevalence, mechanisms, and sex difference of lifetime traumatic brain injury (TBI) in a precariously housed sample. We also examined the impact of TBI severity and timing on becoming and staying homeless. 285 precariously housed participants (adults *n* = 226, youths *n* = 59) completed the Brain Injury Screening Questionnaire (BISQ) in addition to other health assessments. A history of TBI was reported in 82.1% of the sample, with 64.6% reporting > 1 TBI, and 21.4% reporting a moderate or severe TBI (msTBI). 10.1% of adults had traumatically-induced lesions on MRI scans. Assault was the most common mechanism of injury overall, and females reported significantly more TBIs due to physical abuse than males (adjusted OR = 1.26, 95% CI = 1.14 – 1.39, *p* = 9.18e^-6^). The first msTBI was significantly closer to the first experience of homelessness (*b* = 2.79, *p* = 0.003) and precarious housing (*b* = 2.69, *p* = 7.47e^-4^) than was the first mild TBI. Traumatic brain injuries more proximal to the initial loss of stable housing were associated with a longer lifetime duration of homelessness (RR = 1.04, 95% CI = 1.02 – 1.06, *p* = 6.8e^-6^) and precarious housing (RR = 1.03, 95% CI = 1.01 – 1.04, *p* = 5.5e^-10^). These findings demonstrate the high prevalence of TBI in vulnerable persons and the severity- and timing-related risk that TBI may confer for the onset and prolongation of homelessness.

## INTRODUCTION

An estimated 2.5–3.5 million people experience homelessness annually in the USA, with considerably higher estimates worldwide.^1^ Homeless individuals have significantly greater rates of premature mortality, infectious diseases, and psychiatric illness than the general population.^1; 2^ Youth (aged 16–24) make up approximately 20% of the Canadian homeless population.^3^ Homeless and precariously housed youth may have differential risk factors and health outcomes, and represent a uniquely vulnerable demographic within this marginalized population.^4^ Though not formally ‘homeless,’ individuals who live in precarious housing (e.g., shelters, single-room occupancy hotels, rooming houses, etc.) have comparable mortality, chronic health conditions, and unmet health care needs.^5; 6^

Several recent studies have reported that homeless and precariously housed adults and youth have a high (53%–65% adult; 43% youth) lifetime prevalence of traumatic brain injury (TBI).^7-10^ Unlike the general population, these homeless and precariously housed individuals most often sustain TBI through violent mechanisms such as assault.^11; 12^ Moreover, homeless or precariously housed individuals who report a history of TBI are more likely to endorse a history of psychiatric diagnoses and substance abuse issues,^13; 14^ lower physical and mental quality of life,^7; 8^ seizures,^7;15^ and higher rates of emergency department and primary care physician use.^8; 14^

The extent to which TBI is associated with becoming or remaining homeless or precariously housed remains largely unexplored. Cursory examinations of the relationship between TBI and the initial loss of stable housing have indicated that the majority individuals who had a history of TBI reported receiving their first TBI prior to their first episode of homelessness.^7; 10-12; 16; 17^ However, these studies have not considered severity, i.e., whether moderate or severe TBI is more closely associated with the initial loss of stable housing than is mild TBI. Furthermore, no previous studies have evaluated the relationship between the timing of TBI and the lifetime duration of homelessness or precarious housing. Given that TBI can result in long-term disability and both neurologic and psychiatric disorders,^18-20^ TBI closer to becoming homeless or precariously housed may be associated with a prolonged loss of stable housing. Evidence that TBI contributes to the experience of homelessness has important implications for care delivery to this vulnerable population, and for public health policy.

In this study we first sought to characterize the lifetime history of TBI in a precariously housed sample by examining the lifetime prevalence, severity, mechanisms, and sex differences of TBI in this population. We also stratified descriptive analyses by adults (≥ age 30 at assessment) and youth (<29.9 at assessment) to capture differences in the lifetime burden of TBI. Second, we sought to characterize the relationship between both the severity and temporality of TBI with the onset and duration of homelessness and precarious housing. We hypothesized that more severe TBI would occur closer to the initial onset of loss of stable housing. We also hypothesized that TBI more proximal to the initial loss of stable housing would be associated with a longer duration of homelessness or precarious housing.

## METHODS

### Study design and participants

The Hotel Study is an ongoing longitudinal prospective observational study of individuals who live in precarious housing within an impoverished neighbourhood of Vancouver, Canada. Methodology and baseline characteristics for the study sample have been reported previously.^4; 15; 21^ Briefly, between 2008 and 2017, staggered recruitment was used to enroll participants from single-room occupancy hotels and the local downtown community court. Single-room occupancy hotel rooms often fail to meet the minimum housing standards for Canada of being adequate, affordable, and suitable, in that they are often in disrepair, cost more than 30% of total before-tax household income, and house many tenants in facilities with limited space.^22^ Participants were eligible for inclusion if they were 18 years of age or older, able to communicate in English, and able provide written informed consent. Participants enrolled in the Hotel Study receive ongoing monthly follow-up interviews for up to 10 years (e.g., urine drug screens, general medical and symptom questionnaires, housing status), and yearly comprehensive neuropsychological and physician-administered neuropsychiatric examinations, which include detailed assessments of general and psychosocial functioning. Multimodal brain MRI is performed annually or biennially. For descriptive analyses in the present study we evaluate adults (≥ age 30 at assessment) and youth (<29.9 at assessment) separately.^4^ The study was approved by the Clinical Research Ethics Board of the University of British Columbia and the Research Ethics Board at Simon Fraser University in accordance with the Tri-Council Policy and Declaration of Helsinki.

### Demographic and housing measures

Baseline sociodemographic data were collected by trained research assistants using structured interviews at study entry. Psychiatric diagnoses at study entry were assigned by a psychiatrist (WGH, OL) after reviewing all available clinical information with the Best Estimate Clinical Evaluation and Diagnosis II in accordance with the Diagnostic and Statistical Manual for Mental Disorders-TR Fourth Edition criteria.^23; 24^ Housing history prior to study entry was collected as part of the baseline sociodemographic interview. Alterations in housing status were updated monthly during the study. In the present study, *homelessness* was defined as living on the street; in shelters or vehicles; travelling or camping; having no fixed address; or couch surfing. *Precarious housing* was defined as living in a single-room occupancy hotel room, rooming house, or boarding house, according to the Canadian definition of precarious housing.^22^

### Traumatic brain injury assessment and neuroimaging

Lifetime history of TBI was assessed with Part I of the Brain Injury Screening Questionnaire (BISQ).^25^ We classified the severity of TBI using the World Health Organization classification of TBI severity in conjunction with the self-reported duration of loss of consciousness on the BISQ.^26^ Mild TBI was considered to be any self-reported blow to the head, accompanied by loss of consciousness for less than 30 minutes or any period of being dazed and confused. Moderate TBI was considered to be any self-reported blow to the head accompanied by loss of consciousness for between 30 minutes and 24 hours, and severe TBI was considered to be any self-reported blow to the head accompanied by loss of consciousness for more than 24 hours. To evaluate mechanisms of injury, we grouped mechanisms into categories of motor vehicle accidents or being struck as a pedestrian; falling or being struck by an object; sports and recreation; falling due to drug or alcohol blackout; being assaulted or mugged; and physical abuse.

Hotel Study participants complete yearly or biennial MRI scans on a Philips 3.0 Tesla Achieva scanner with an eight-channel head coil (Philips Healthcare, Amsterdam, Netherlands). For the present study we assessed high-resolution T1-weighted isotropic 3D IR-weighted fast-field echo anatomic images. Specific parameters were TE = 3.5 ms, TR = 8.1 ms, flip angle = 8, FOV = 256 mm × 256 mm, acquisition matrix = 256 × 250, reconstruction matrix = 256 × 256, voxel size = 1.0 × 1.0 × 1.0 mm^3^, 190 contiguous slices, gap = 0, scan duration = 443 s, SENSE = 1, no partial imaging acceleration. All scans were reviewed by certified neuroradiologists (ATV, MKH) for clinically relevant incidental findings. We excluded scans that were incomplete or contained significant motion artifact. In the present study we report the prevalence of lesions seen on T1-weighted MRI scans that were deemed to be most likely traumatically-induced by consensus of a three-member team comprised of a neuroradiologists and a qualified neuroscientist (DJL, ATV, or MKH), a TBI neurologist (WJP) and a stroke neurologist (TSF).

### Statistical analysis

We compared demographic characteristics and TBI history variables between adults and youth using independent-samples *t*-tests, Mann-Whitney *U* tests, Chi-squared tests with Yates’ continuity corrections, or Fisher’s Exact Tests as appropriate. All other analyses were conducted on the full sample including both adult and youth, adjusted for age. We evaluated sex differences across mechanism of TBI using logistic regression models with the mechanism of interest as the dependent variable, adjusting for age. To evaluate whether severity of worst lifetime TBI was associated with the age of first loss of stable housing, we used multiple linear regression models to evaluate whether history of TBI (no TBI, only mild TBI, or moderate or severe TBI) was associated with the age at first homelessness or precarious housing, adjusted for sex. To assess whether moderate or severe TBI was more closely associated with the initial loss of stable housing, we used multiple linear regression models to compare the mean number of years between the first most severe TBI (i.e. the first mild TBI in those whose worst TBI was mild, or the first moderate or severe TBI in those whose worst TBI was moderate or severe) and the first experience of homelessness and precarious housing, adjusted for age and sex. Finally, we evaluated whether fewer number of years between the first TBI and the first experience of homelessness or precarious housing was associated with a longer lifetime duration of homelessness or precarious housing using negative binomial regression, adjusted for age and sex. Exponentiating coefficients from the negative binomial regression resulted in rate ratios and their 95% confidence intervals. All analyses were performed in *R* version 3.6.0.^27^

## RESULTS

### Demographic characteristics

Our study sample consisted of 285 participants (adult sample *n=*226; youth sample *n*=59) who had completed the BISQ, and demographic and psychiatric diagnoses are reported in Table 1. The adult sample was significantly older and had experienced a longer lifetime duration in precarious housing than the youth sample. When evaluated as a rate (lifetime duration divided by age) the youth sample had experienced a greater proportion of their lifetime spent in homelessness (*U*=5044.5, *p*=0.005). Those in the youth sample became homeless and precariously housed at a significantly younger age than those in the adult sample. At entry into the study, the youth sample had a higher prevalence of psychotic disorders, methamphetamine dependence, and cannabis dependence than the adult sample, while the adult sample had a higher prevalence of opioid and cocaine dependence than the youth sample.

**Table 1.** Demographic and clinical characteristics for participants, further stratified into the adult and youth samples.

|  | Full sample |  |  | Adult |  |  | Youth |  |  | Test statistic | *p*-value |
| --- | --- | --- | --- | --- | --- | --- | --- | --- | --- | --- | --- |
| Demographic | N | Median | IQR | N | Median | IQR | N | Median | IQR |  |  |
| Age | 285 | 45 | 20 | 226 | 48 | 12.3 | 59 | 24 | 4 | N/A<sup>a</sup> | N/A |
| Monthly income (CAD) | 280 | 758 | 395 | 224 | 766.5 | 429 | 56 | 820 | 410 | 5836 | 0.394 |
| Age at first homelessness | 211 | 23 | 15.5 | 158 | 27 | 17 | 53 | 19 | 4 | 1979 | 9.19e<sup>-09</sup> |
| Age at first precarious housing | 266 | 29 | 18 | 209 | 34 | 16 | 57 | 21 | 3 | 1972.5 | 9.73e<sup>-15</sup> |
| Months spent homeless | 283 | 18.2 | 52 | 224 | 17.5 | 58.1 | 59 | 19.3 | 40.4 | 6914.5 | 0.582 |
| Months spent in precarious housing | 266 | 48 | 92.3 | 209 | 68 | 102 | 57 | 14.2 | 30 | 2756.5 | 5.14e<sup>-10</sup> |
|  | *n* | **N** | **%** | *n* | **N** | **%** | *n* | **N** | **%** | **Test stat** | ***p*-value** |
| Female | 63 | 285 | 22.1 | 48 | 226 | 21.2 | 15 | 59 | 25.4 | 0.2638 | 0.608 |
| Completed high school | 72 | 282 | 25.5 | 54 | 223 | 24.2 | 19 | 59 | 32.2 | 1.1633 | 0.281 |
| Obtained GED | 43 | 282 | 15.2 | 41 | 223 | 18.4 | 2 | 59 | 3.39 | 6.9997 | 0.008 |
| Obtained trade cert., diploma, or degree | 81 | 275 | 29.5 | 67 | 219 | 30.6 | 14 | 56 | 25 | 0.4293 | 0.512 |
| **Diagnoses at study entry** | *n* | **N** | **%** | *n* | **N** | **%** | *n* | **N** | **%** | **Test stat** | ***p*-value** |
| Psychotic disorder | 135 | 285 | 47.4 | 100 | 226 | 44.2 | 35 | 59 | 59.3 | 4.171 | 0.041 |
| Mood disorder | 80 | 285 | 28.1 | 61 | 226 | 27 | 19 | 59 | 32.2 | 0.3978 | 0.528 |
| Alcohol | 53 | 285 | 18.6 | 39 | 226 | 17.3 | 14 | 59 | 23.7 | 0.90238 | 0.342 |
| Methamphetamine | 88 | 285 | 30.9 | 62 | 226 | 27.4 | 26 | 59 | 44.1 | 5.311 | 0.021 |
| Cocaine | 160 | 285 | 56.1 | 156 | 226 | 69 | 4 | 59 | 6.78 | 71.116 | 3.37e<sup>-17</sup> |
| Opioid | 148 | 285 | 51.9 | 128 | 226 | 56.6 | 20 | 59 | 33.9 | 8.801 | 0.003 |
| Cannabis | 109 | 285 | 38.2 | 70 | 226 | 31 | 39 | 59 | 66.1 | 22.98 | 1.64e<sup>-06</sup> |
| *Note:* Test statistics are for the difference in medians or proportions between the adult and youth cohorts; <sup>a</sup>As adults and youth were classified based on age we did not compare age statistically. |  |  |  |  |  |  |  |  |  |  |  |

### Characteristics of lifetime TBI

Characteristics of lifetime TBI are reported in Table 2. The median age at first TBI in the full sample was 12, and the modal age was six. The youth sample reported a significantly younger median age at first TBI (*U*=3802, *p*=0.001), and a trend towards younger median age at first moderate or severe TBI than the adult sample (*U*=95, *p*=0.059).

**Table 2.**
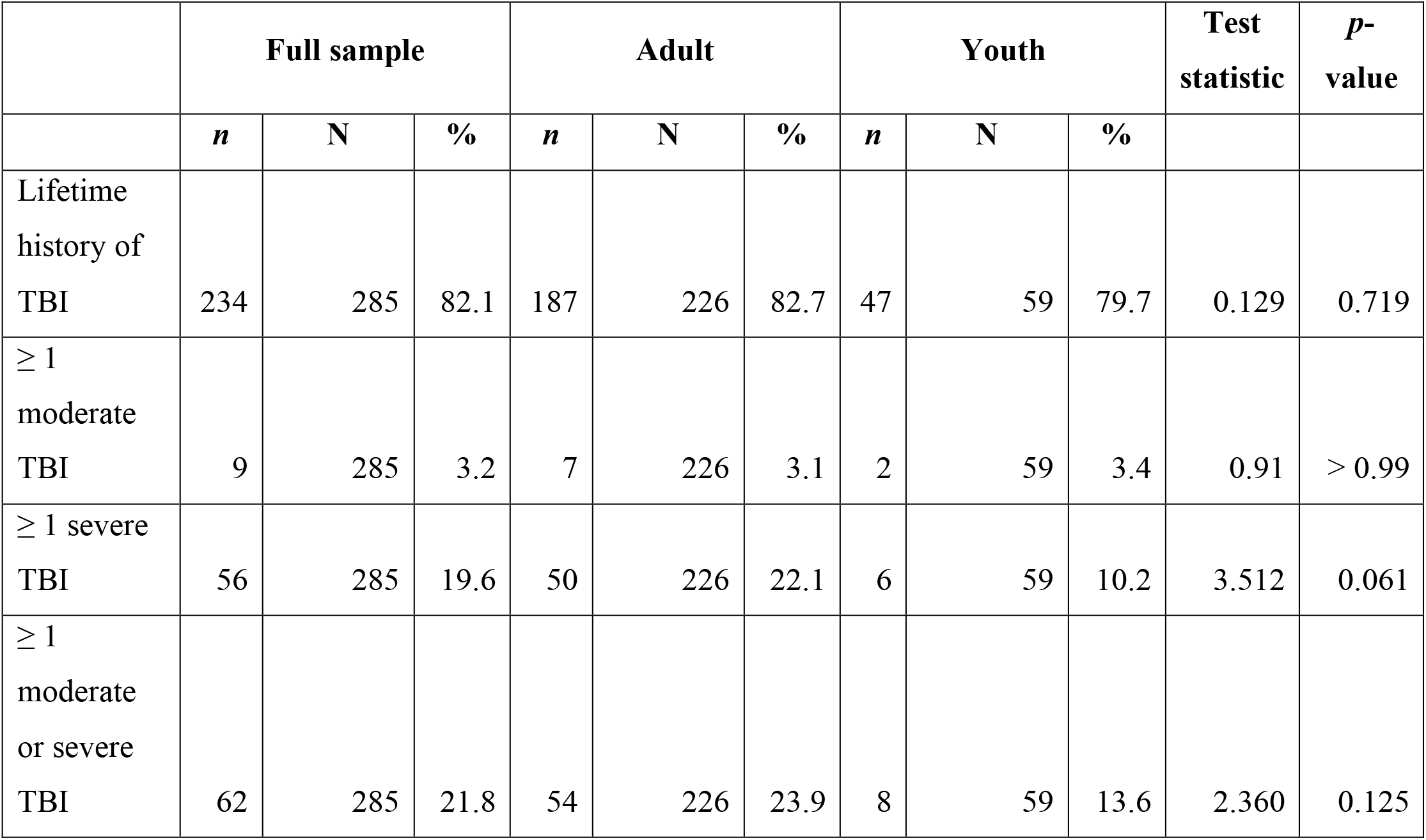
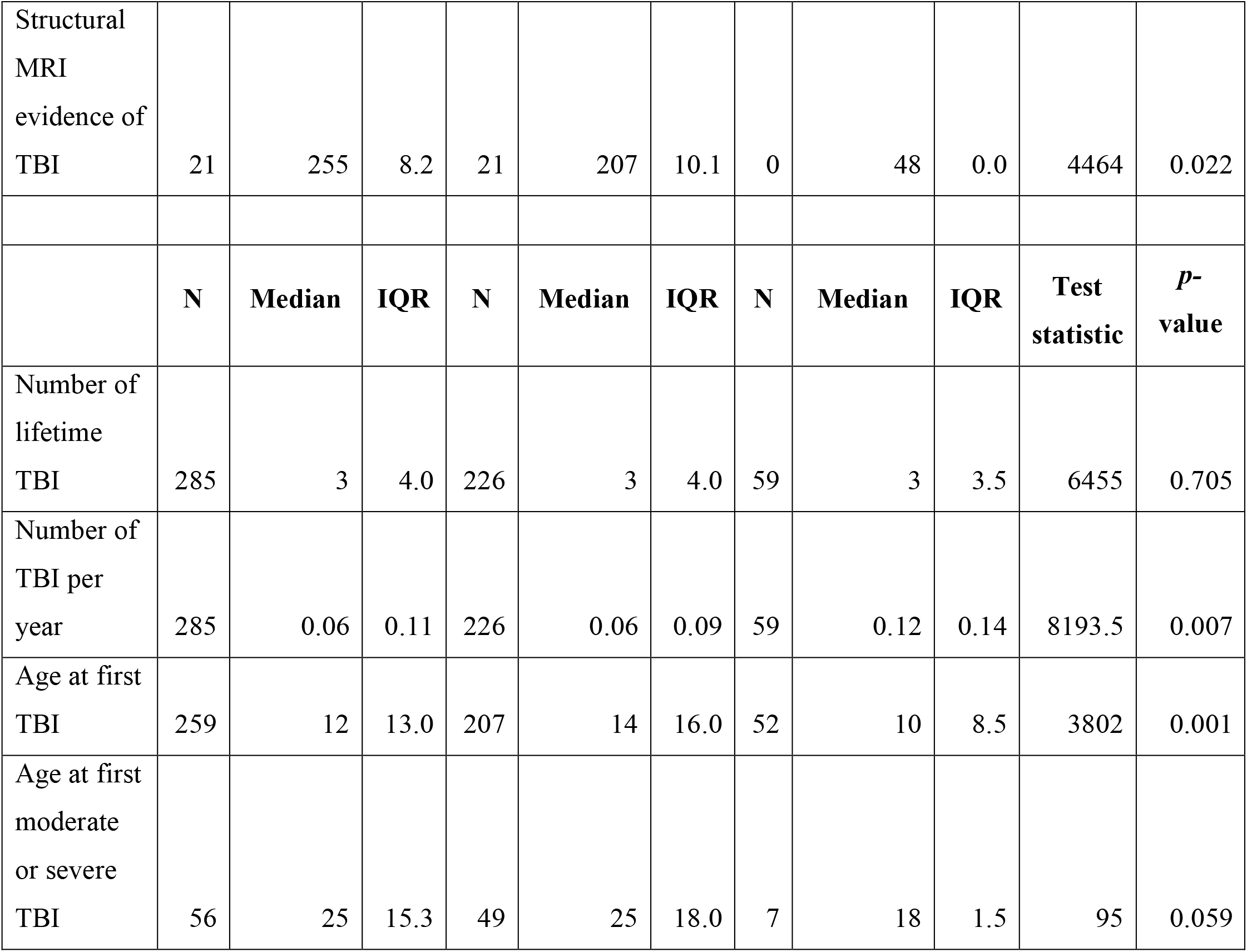
Characteristics of lifetime history of TBI in the full sample, and both the adult and youth samples.

|  | Full sample |  |  | Adult |  |  | Youth |  |  | Test statistic | *p*-value |
| --- | --- | --- | --- | --- | --- | --- | --- | --- | --- | --- | --- |
|  | *n* | N | % | *n* | N | % | *n* | N | % |  |  |
| Lifetime history of TBI | 234 | 285 | 82.1 | 187 | 226 | 82.7 | 47 | 59 | 79.7 | 0.129 | 0.719 |
| ≥ 1 moderate TBI | 9 | 285 | 3.2 | 7 | 226 | 3.1 | 2 | 59 | 3.4 | 0.91 | > 0.99 |
| ≥ 1 severe TBI | 56 | 285 | 19.6 | 50 | 226 | 22.1 | 6 | 59 | 10.2 | 3.512 | 0.061 |
| ≥ 1 moderate or severe TBI | 62 | 285 | 21.8 | 54 | 226 | 23.9 | 8 | 59 | 13.6 | 2.360 | 0.125 |
| Structural MRI evidence of TBI | 21 | 255 | 8.2 | 21 | 207 | 10.1 | 0 | 48 | 0.0 | 4464 | 0.022 |
|  | **N** | **Median** | **IQR** | **N** | **Median** | **IQR** | **N** | **Median** | **IQR** | **Test statistic** | ***p*-value** |
| Number of lifetime TBI | 285 | 3 | 4.0 | 226 | 3 | 4.0 | 59 | 3 | 3.5 | 6455 | 0.705 |
| Number of TBI per year | 285 | 0.06 | 0.11 | 226 | 0.06 | 0.09 | 59 | 0.12 | 0.14 | 8193.5 | 0.007 |
| Age at first TBI | 259 | 12 | 13.0 | 207 | 14 | 16.0 | 52 | 10 | 8.5 | 3802 | 0.001 |
| Age at first moderate or severe TBI | 56 | 25 | 15.3 | 49 | 25 | 18.0 | 7 | 18 | 1.5 | 95 | 0.059 |

Eighty two percent of the study sample reported at least one lifetime TBI, and the proportion of adults and youth who endorsed a history of TBI was not significantly different. Sixty-four percent of participants reported more than one TBI in their lifetime and 59.6% of participants reported a TBI that resulted in loss of consciousness. The median number of lifetime TBI (median=3) did not differ between the adults and youth. In contrast, when analyzed as a rate (number of TBI per year), the youth sample reported a significantly higher number of TBI per year (*U*=8193.5, *p*=0.007). In this sample, 21.8% reported a moderate or severe TBI, with a higher, though not statistically significant, prevalence in the adult sample than the youth sample. Individuals whose worst TBI was moderate or severe also reported a higher number of lifetime TBIs (*U*=3933, *p*=0.002). All participants who had T1-weighted MRI evidence of prior TBI (10.1%) were in the adult sample. In total, 26.7% of all participants in this sample reported either a history of moderate or severe TBI, or had evidence of TBI visible on neuroimaging.

There were no significant differences in mechanism of TBI between the adult and youth sample, and thus, the mechanisms by which participants in the full sample received one or multiple TBIs is shown in Figure 1. Being assaulted or mugged and falling were the most commonly reported mechanisms of injury. There was a trend towards a higher proportion of males reporting more TBI through being assaulted or mugged than females (adjusted OR=1.13, 95% CI=0.99–1.31, *p*=0.070). Females reported significantly more TBIs through physical abuse as compared to males (Figure 1 inset; adjusted OR=1.26, 95% CI=1.14–1.39, *p*=9.18e^-06^).

**Figure 1.**
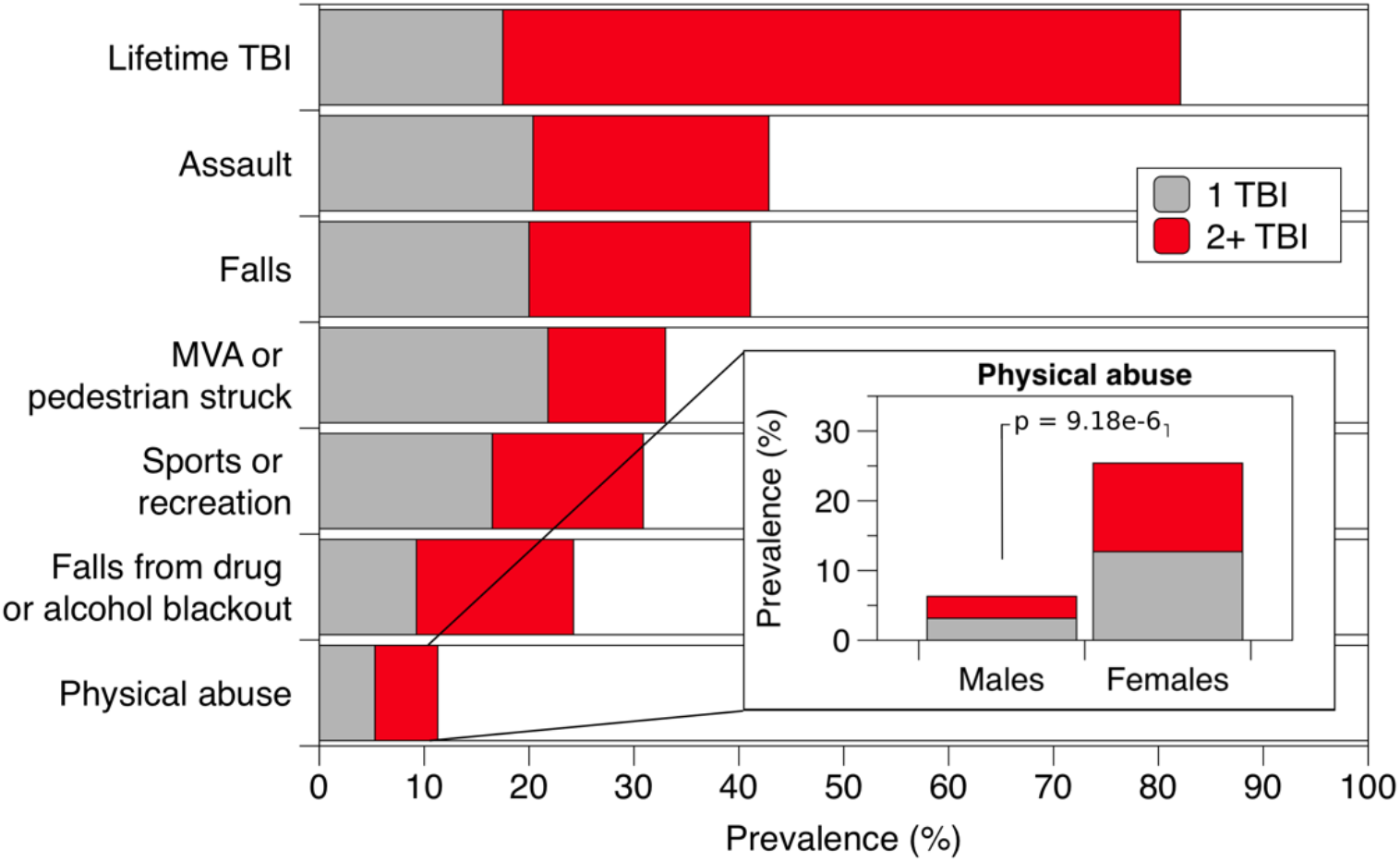
Lifetime prevalence of single and multiple TBIs by mechanism. *Note*: The ‘Falls’, ‘MVA or pedestrian struck’, and ‘Sports and recreation’ categories encompass multiple discrete mechanisms on the BISQ.

### Relationship between severity of TBI and losing stable housing

To evaluate whether history or severity of TBI was associated with earlier loss of stable housing, we compared the mean age of first homelessness or precarious housing between those with no history of TBI, those whose worst TBI was mild, and those whose worst TBI was moderate or severe. Age at first homelessness and precarious housing stratified by the severity of worst TBI is shown in Figure 2a-b. Neither a history of mild TBI (*b*=-0.89, *p*=0.853) nor a history of moderate or severe TBI (*b*=-0.53, *p*=0.794) were associated with an earlier age at first homelessness, as compared to those who did not report TBI, adjusted for age and sex. Findings were similar for age at first precarious housing (no TBI=reference; mild TBI: *b*=-2.15, *p*=0.146; moderate or severe TBI: *b*=-2.03, *p*=0.245), adjusted for age and sex.

**Figure 2.**
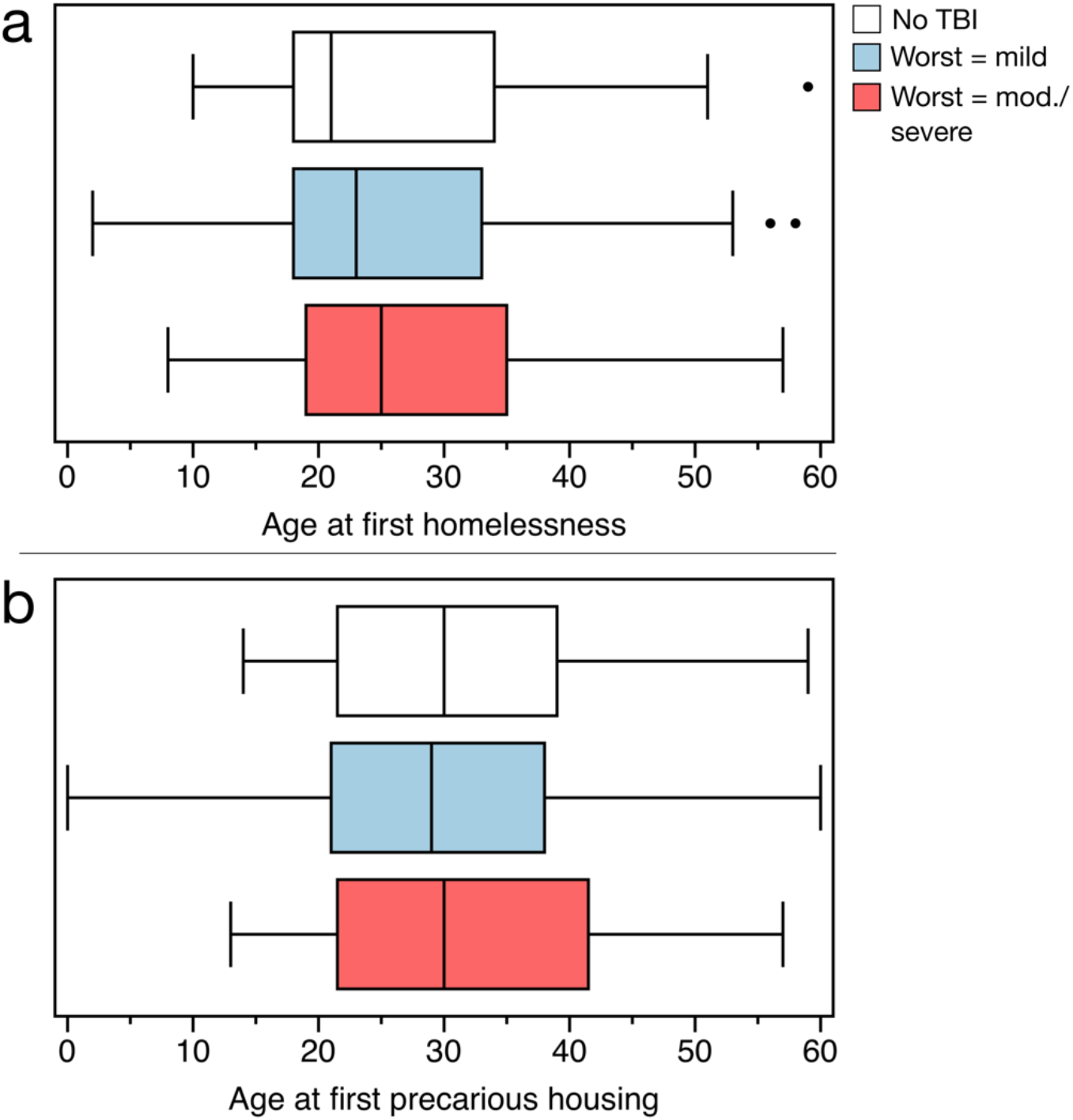
Age at first homelessness (a) and age at first precarious housing (b) for the full sample who had no TBI (white), whose worst TBI was mild (blue) and whose worst TBI was moderate or severe (red). Boxes represent interquartile range, whiskers denote 1.5 standard deviations, and dots represent outliers more than 1.5 standard deviations from the median.

We then evaluated the time interval between the participants’ most severe TBI and the onset of homelessness or precarious housing, adjusted for age. The distributions of the number of years between the first most severe TBI and the onset of homelessness and precarious housing are shown in Figure 3a-b. For adults with a history of only mild TBI, the median number of years between the first mild TBI and the first experience of homelessness was 13 years and for precarious housing was 17.5 years. For adults whose worst TBI was moderate or severe, the median number of years between their first moderate or severe TBI and the first experience of homelessness was zero years (i.e., the first moderate or severe TBI occurred in the same year as the first experience of homelessness) and for precarious housing was eight years.

**Figure 3.**
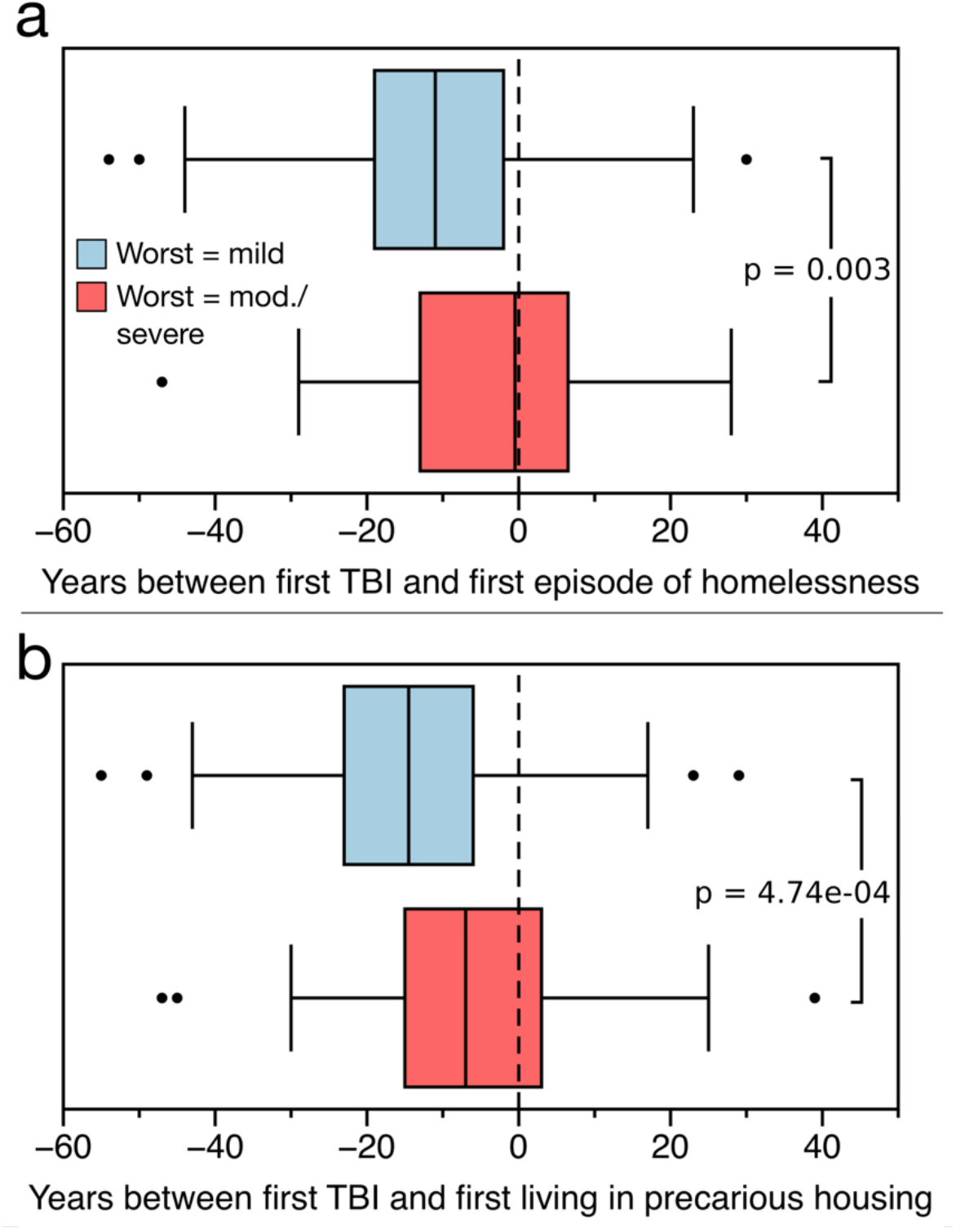
Difference in the number of years between the first mild TBI (blue) or the first moderate/severe TBI (red) and the first experience of homelessness (a) and the first experience of precarious housing (b). Negative values denote a TBI occurring before the first experience of homelessness or precarious housing. Boxes represent interquartile range, whiskers denote 1.5 standard deviations, and dots represent outliers more than 1.5 standard deviations from the median.

Findings were similar in the youth sample: In youths with a history of only mild TBI, the median number of years between the first mild TBI and the first experience of homelessness was 6.5 years and for precarious housing was 10.5 years. In youths with a history of moderate or severe TBI, the median number of years between the first moderate or severe TBI and the first experience of homelessness was 0.5 years and for precarious housing was 3 years. In the full sample, the first moderate or severe TBI occurred significantly closer to the year of first homelessness (*b*=2.79, *p*=0.003), adjusting for age and sex. Similarly, we found that the first moderate or severe TBI occurred significantly closer to the year of first precarious housing (*b*=2.69, *p*=7.47e^-4^), adjusting for age and sex.

### Relationship between the timing of TBI and remaining homeless or precariously housed

Finally, we used negative binomial regression to evaluate whether TBI closer to the initial loss of stable housing was associated with a longer lifetime duration of homelessness or precarious housing. In this sample, 58.2% of participants reported that they received their first TBI prior to first becoming homeless, and 74.0% reported that the received their first TBI prior to becoming precariously housed. Traumatic brain injury closer to the first experience of homelessness was significantly associated with a longer lifetime duration of homelessness, adjusting for age and sex (RR=1.04, 95% CI=1.02–1.06, *p*=6.8e^-6^). Thus, for every additional year closer to becoming homeless that participants received their first TBI, there was a mean increase of 4% in their lifetime duration of homelessness. The results were similar for the relationship between the first TBI and the first experience of precarious housing (RR=1.03, 95% CI=1.01–1.04, *p*=5.5e^-10^), adjusted for age and sex.

## DISCUSSION

In this sample of precariously housed adults and youth, 82.1% reported a lifetime history of TBI and 21.7% reported a history of moderate or severe TBI. One previous meta-analysis (*n*=25,134) found that 12% of the general population reported a lifetime history of TBI.^28^ A more recent state-wide survey (in Ohio, USA; *n*=6,998) estimated that 21.7% of the general population had at least one TBI with loss of consciousness and 2.6% had experienced at least one moderate or severe TBI.^29^ In comparison, the lifetime prevalence of TBI in this precariously housed sample is 3.8–6.8 times that of the general population. Additionally, the lifetime prevalence of moderate or severe TBI in this population is nearly 10 times that of estimates in the general population. Notably, 8.2% of participants in this sample (adults and youth combined) had evidence of trauma visible on T1-weighted MRI scans.

Homeless and precariously housed youth in this study reported a disproportionately high burden of TBI; those in the youth sample sustained their first TBI at a significantly younger age and sustained more TBIs per year as compared with the adult sample. Both adults and youth generally reported that their first TBI occurred early in life. While the ages at first TBI and first moderate or severe TBI in the adult sample were similar to those reported in the general population,^30^ the ages at first TBI in the youth sample were lower than those in the general population. Traumatic brain injury in the critical developmental period of adolescence is a risk factor for the development of schizophrenia spectrum disorders, bipolar disorder, and other organic mental illnesses.^31^ While we did not evaluate TBI as a risk factor for the onset of diagnosed psychiatric illness, there was a high prevalence of both TBI and diagnosed psychiatric illness in this sample. Future studies should evaluate the impact of TBI in exacerbating existing psychiatric illness in vulnerable populations. Traumatic brain injury is also associated – in a dose-response relationship – with higher risk for low educational attainment, receiving disability pension, and welfare recipiency in adulthood.^30^ Thus, TBI may be an important factor in the broad range of challenges faced by homeless and precariously housed populations.

The most commonly reported mechanism of injury in this sample was assault, corroborating previous studies that evaluated mechanism of injury in this population.^12; 16; 32; 33^ Conversely, in the general population, sustaining TBI through assault is not a predominant cause of TBI causing hospitalization.^34^ Females had a significantly higher odds of sustaining TBI through physical abuse, an important and actionable finding that parallels recent studies that have evaluated TBI in individuals who are survivors of intimate partner violence.^35^

Our findings also highlight notable associations between the severity and timing of TBI with housing status. First, moderate or severe TBI was closely related to the initial loss of stable housing, which suggests that a considerable proportion of newly homeless and precariously housed individuals may be acutely experiencing sequelae related to TBI. Second, TBI closer to the initial loss of losing stable housing was associated with a longer duration of homelessness and precarious housing, further suggesting that sequelae of TBI may be an important mediator of functional capacity in those who have transitioned to homelessness, and may create additional barriers to exiting homelessness. Importantly, the relationship between TBI and homelessness is likely bidirectional given that homelessness is itself associated with a greater risk of incident TBIs, whether through an increase in violent encounters, substance abuse, or other factors.^13; 36^

This study has several strengths and limitations. First, we assessed lifetime TBI using the BISQ, which is a screening tool designed specifically to characterize lifetime TBI, and validated in similar populations.^25^ Many previous studies that have assessed TBI in homeless and precariously housed populations have used only a single question (i.e. ‘Have you ever experienced a blow to the head…’) to ascertain TBI history, yet, single-item screening questions for head injury have been shown to considerably underestimate the prevalence estimated by more comprehensive methods.^37; 38^ Thus, our estimate of lifetime prevalence may be a more representative estimate of the true lifetime prevalence of TBI in this population. However, TBI history was still largely assessed through participant self-report. Relying on self-reported loss of consciousness is a necessary but potentially unreliable method of retrospectively determining TBI severity.^39^ Additionally, we were not able to identify the specific year of every injury after the first TBI, which precluded a more detailed analysis. Finally, participants with significant comorbid substance use disorders or economic disadvantage may be missed or lost to follow-up in longitudinal studies.^40; 41^ While the Hotel Study is focused specifically on this vulnerable population, the study necessitates coordinating and attending follow-up visits, and thus we may have inadvertently missed individuals who had the most severe manifestations of psychiatric illness (and possibly those with a higher lifetime burden of TBI).

We found that a majority of homeless and precariously housed individuals reported a lifetime history of TBI, and more than one-fifth reported a history of moderate or severe TBI. Youth in this sample experienced a disproportionately high burden of TBI, including more TBIs per year and a younger first age at initial TBI. Injuries for both adults and youth were most commonly sustained through assaults and falls, though females were more vulnerable to sustaining TBI through physical abuse. We showed that moderate or severe TBIs generally occurred closer to the initial loss of stable housing, and that sustaining TBI closer to the initial loss of stable housing was associated with a longer lifetime duration of both homelessness and precarious housing. Notably, in both our analyses of severity and temporality, we found comparable results for homelessness and precarious housing. This highlights the notion that despite being ‘housed’, individuals living in precarious housing share largely similar characteristics and consequences as homeless individuals. Future research that uses standardized assessment methods in prospective study designs is needed to better understand the full scope of the impact of TBI in this population.

## Data Availability

Data from this study may be sent to interested authors upon reasonable request.

## ACKNOWLEDGEMENTS

We thank the many research assistants and volunteers who are integral to this study.

## DECALRATION OF CONFLICTING INTERESTS

Dr. Honer has received consulting fees or sat on paid advisory boards for: the Canadian Agency for Drugs and Technology in Health, AlphaSights, Guidepoint, In Silico, Translational Life Sciences, Otsuka, Lundbeck, and Newron. Dr. Panenka is the founder and CEO of Translational Life Sciences, an early stage biotechnology company. He is also on the scientific advisory board of Medipure Pharmaceuticals and Vitality Biopharma, and in the past has been on the board of directors for Abbatis bioceuticals and on the advisory board of Vinergy Resources. All of these companies are early stage biotechnology enterprises with no relation to brain injury. The other authors have no competing financial interests.

